# Epidemic Progression and Vaccination in a Heterogeneous Population. Application to the Covid-19 epidemic

**DOI:** 10.1101/2020.12.06.20244731

**Authors:** Vitaly Volpert, Malay Banerjee, Swarnali Sharma

**Affiliations:** Institut Camille Jordan, UMR 5208 CNRS, University Lyon 1, 69622 Villeurbanne, France; INRIA Team Dracula, INRIA Lyon La Doua, 69603 Villeurbanne, France; Peoples Friendship University of Russia (RUDN University) 6 Miklukho-Maklaya St, Moscow, 117198, Russian Federation; Department of Mathematics & Statistics, IIT Kanpur, Kanpur - 208016, India; Department of Mathematics, Vijaygarh Jyotish Ray College, Kolkata - 700032, India

**Keywords:** Covid-19, heterogeneous population, final size, duration of epidemic, vaccination

## Abstract

The paper is devoted to a compartmental epidemiological model of infection progression in a heterogeneous population which consists of two groups with high disease transmission (HT) and low disease transmission (LT) potentials. Final size and duration of epidemic, the total and current maximal number of infected individuals are estimated depending on the structure of the population. It is shown that with the same basic reproduction number *R*_0_ in the beginning of epidemic, its further progression depends on the ratio between the two groups. Therefore, fitting the data in the beginning of epidemic and the determination of *R*_0_ are not sufficient to predict its long time behaviour. Available data on the Covid-19 epidemic allows the estimation of the proportion of the HT and LT groups. Estimated structure of the population is used for the investigation of the influence of vaccination on further epidemic development. The result of vaccination strongly depends on the proportion of vaccinated individuals between the two groups. Vaccination of the HT group acts to stop the epidemic and essentially decreases the total number of infected individuals at the end of epidemic and the current maximal number of infected individuals while vaccination of the LT group only acts to protect vaccinated individuals from further infection.

## 1 Introduction

Covid-19 epidemic has stimulated an unprecedented interest to the epidemiological models, mostly, compartmental ODE models. There are numerous recent works devoted to fitting the available data, calculating the basic reproduction number, and making predictions about the further epidemic progression (see [1, 2, 3, 4, 5, 6, 7] and the references therein). These models give a good description of the evolution of the number of infected individuals and the sizes of other classes involved with the epidemiological models in the beginning of epidemic, and they take into account the influence of the measures of social distancing and some other meausres to prevent the rapid epidemic spread. The situation is more complex with the prediction of the future epidemic progression because the parameters of the models are influenced by the measures of social distancing and other behavarial changes, and hence it is impossible to predict various scenario in advance.

At the end of the first year of the epidemic and during its second wave, sufficient amount of data are available to model the long time epidemic progression, including the development of collective immunity, the final size of epidemic, and the influence of vaccination on further epidemic growth profile. An important assumption here is that recovered and vaccinated individuals do not become susceptible any more. Though it is one of the most important open questions of the coronavirus disease, and immunological studies show that the quantity of antibodies in recovered individuals can be highly variable [8], we will adopt here this hypothesis.

The influence of heterogeneity of the population with respect to its role in the epidemic progression is largely discussed in the existing literature [9, 10, 11, 12, 13]. Different age and social groups can have different frequency of interactions and implementation of the measures of social distancing. Furthermore, the so-called superspreaders, a relatively small group of people with a large number of social interactions, play an important role in the coronavirus epidemic [14]. Consideration of two types of individuals, one having frequent social interaction and the other having restricted/cautious social interaction, together in a single group with an average infectivity can lead to erroneous predictions.

In this work we will study how the heterogeneity of the population influences its long time progression including the final size and duration of epidemic. In order to simplify the model and the interpretation of the results, we will consider only two groups in the population, one of them with high disease transmission (HT) and another one with low disease transmission (LT) potentials. If the total size of the population is *N*, the initial size of the first group (HT) is *N*_1_ and the second group (LT) is *N*_2_ such that *N* = *N*_1_ + *N*_2_, then we introduce the coefficient *k* = *N*_1_*/N* characterizing the structure of the population. If *k* = 1, then the whole population belongs to the first group, if *k* = 0, to the second group. In general, *k* adopts the values between 0 and 1. Two extreme values of *k* correspond to a single group epidemic.

The proportion between these two groups strongly influences the final size of epidemic (final number of susceptible individuals *S*_*f*_) (cf. [15]) and the maximal current number of infected individuals *I*_*m*_. This second parameter (*I*_*m*_) is particularly important for the estimation of the necessary number of hospital beds to handle the worst case scenario. It appears that for the same basic reproduction number *R*_0_, the values *S*_*f*_ and *I*_*m*_ can differ several times depending on the parameter *k*. It is important to stress here that fitting the same data in the beginning of epidemic can be done for any value of *k* but further epidemic progression will crucially depend on it. Thus, the initial growth rate does not allow the prediction of long time epidemic progression in a heterogeneous population.

We suggest a method to estimate the structure of the population in each given country during the Covid-19 epidemic on the basis of the available data before, during and after the first lockdown. Since there were no measures of social distancing (obligation of wearing masks, restriction on social gathering, etc.) before the lockdown, we assume that the whole population belonged to the first group (HT), and *k* = 1. On the other hand, during the lockdown, we assume that the whole population respected strict measures of social distancing and other restrictions, and *N*_2_ = *N*, that is *k* = 0. In both cases, we fit the data and determine the parameters of the epidemiological model. After the lockdown when the restrictions are gradually lifted, the population splits into two groups, HT and LT, in certain proportion. We assign the first group the same parameters as before lockdown, and the second group the same parameters as during lockdown. Hence, we have all parameters characterizing each group, and one free parameter *k* which determines the proportion between the groups. The data after lockdown allow us to determine the value of this parameter and to characterize the heterogeneity of the population. Carrying out this analysis for several countries, we obtain *k*≈ 0.1 during the summer period and *k*≈ 0.2 in September-November 2020. This increase of the parameter *k* corresponds to the second wave of the epidemic. The rate of epidemic growth and the size of the second wave are determined by the value of *k*.

Clearly, suggested approach does not take into account the heterogeneity inside each group, possible exchange between the groups, and some other factors. However, it gives a single efficient parameter characterizing the structure of the population and the epidemic progression. We will call this parameter the coefficient of social interaction since *k* = 0 corresponds to low interaction during the lockdown and *k* = 1 to high interaction before the lockdown. Once the lockdown is relaxed, it is expected that people continue to follow some restrictions, either imposed by public authorities or self-imposed. However, different social and professional groups can have different levels of implementation of these restrictions and of the intensity of social contacts. As a result, the population splits into two groups, and the parameter *k* adopts some intermediate value between 0 and 1.

Having determined the structure of the heterogeneous population, we can study the influence of vaccination on the further epidemic progression. The results of the vaccination strongly depend on whether it is applied to HT group or to LT group. In particular, with only 5% of vaccinated individuals (of the whole population) for *k* = 0.2, the total number of infected individuals at the end of epidemic is almost 3 times less than without vaccination, if vaccination is applied to the HT group. If vaccination is applied to the LT group, the effect of vaccination is weak. Hence, vaccination of the first group acts to stop the epidemic while vaccination of the second group only protects vaccinated individuals. Though this result can be expected, the difference in the results of vaccination is quite striking.

The contents of the paper are as follows. In the next section, we introduce and study a model problem of a heterogeneous population. We determine various parameters of epidemic progression and show that with the same initial growth rate, its outcome can strongly differ depending on the structure of the population. In Section 3 we introduce a more complete epidemiological model of heterogeneous population. We apply it to Covid-19 in Section 4 in order to determine the structure of the heterogeneous populations and to model the influence of vaccination on the epidemic progression. Finally, discussion of the model and of the result is presented in Section 5.

## 2 Model problem

We begin the study of epidemic progression in a heterogeneous population with the model problem consisting of susceptible and infected individuals with two sub-populations:

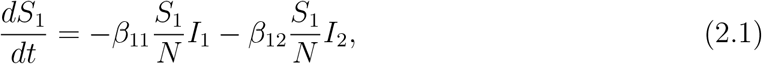

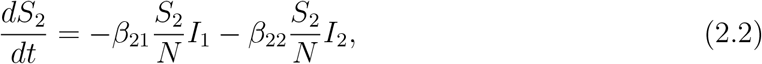

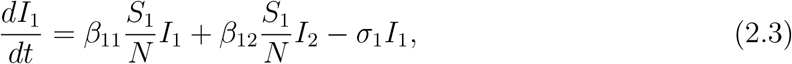

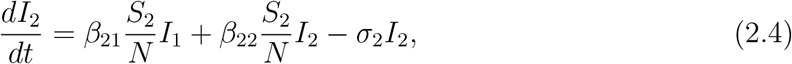

where *β*_*ij*_ are the rates of disease transmissions, *σ*_*j*_ are the clearance rates, and *N* is the total population. This model is similar to the model recently considered in [15]. We will present a more detailed analysis compared to the previous one. Along with basic reproduction number and the final size of epidemic, we will determine the maximal number of infected and will show that for the same value of basic reproduction number, the populations can strongly differ by their final size and the maximum of infected individuals. This effect occurs because of the heterogeneity of the population. Furthermore, we will study the influence of vaccination on the heterogeneous population.

### 2.1 Basic reproduction number

In the beginning of epidemic, *S*_1_ and *S*_2_ can be considered as constant. We set:

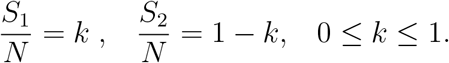

The linearized matrix of the system (2.1) - (2.4) evaluated at disease free equilibrium point (*kN*, (1 − *k*)*N*, 0, 0) is given by

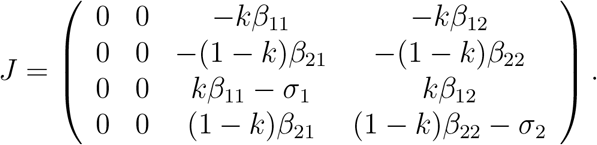

The non-zero eigenvalues of *J* can be obtained from the block matrix

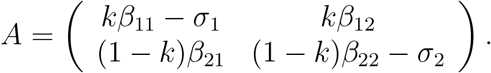

We find maximal eigenvalues of *J* from the equation:

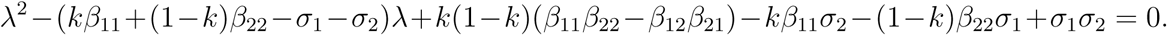

In order to simplify further calculations, we suppose that *σ*_1_ = *σ*_2_ = *σ, β*_12_ = *β*_21_ = (*β*_11_ + *β*_22_)*/*2. Then

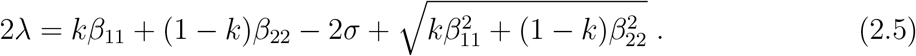

The basic reproduction number *R*_0_ is as follows:

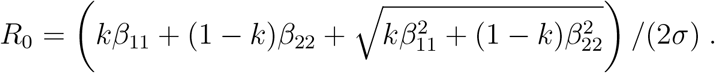

If, moreover, *β*_11_ = *β*_22_ = *β* for some *β*, then *λ* = *β* − *σ, R*_0_ = *β/σ*.

### 2.2 Final size of epidemic

Taking a sum of equations (2.1), (2.3) and (2.2), (2.4), we obtain the equalities:

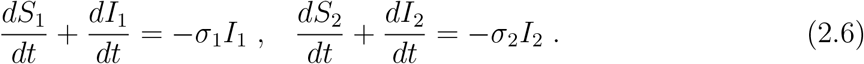

Integrating them from 0 ∞ to and assuming that *I*_*i*_(0) = *I*_*i*_(∞) = 0, *i* = 1, 2, we conclude that

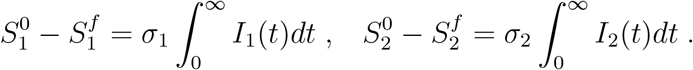

Next, we divide equation (2.1) by *S*_1_, equation (2.2) by *S*_2_ and integrate from 0 to ∞:

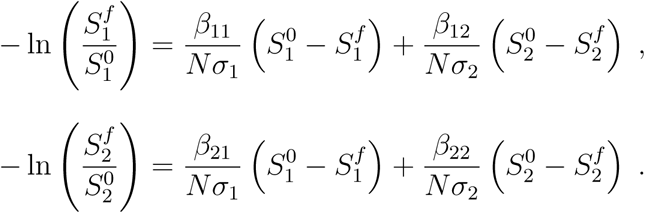

With the notation 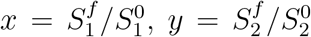, and assumptions *σ*_1_ = *σ*_2_ = *σ, β*_12_ = *β*_21_ = (*β*_11_ + *β*_22_)*/*2, we obtain the system of equations

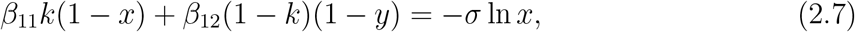

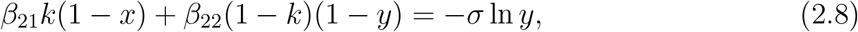

with respect to *x* and *y*. If *β*_11_ = *β*_22_ = *β*, then this system is reduced to the single equation *R*_0_(1 − *x*)+ln *x* = 0 independent of *k*. Its solution gives the final size of susceptible population for the homogeneous population.

In the general case, the solution of this system depends on *β*_11_, *β*_22_, and *k*. We will vary their values in such a way that the basic reproduction number does not change, and we will analyze the final size of epidemic.

Consider the following example: *β* = 2.5, *σ* = 1. Then *λ* = 1.5, *R*_0_ = 2.5. For different values *β*_11_ and *β*_22_ such that (*β*_11_ + *β*_22_)*/*2 = *β*, we find from (2.5) the value of *k* for which *λ* = 1.5:

|  |  |  |  |  |  |  |  |
| --- | --- | --- | --- | --- | --- | --- | --- |
| $\beta_{11}$ | 2.6 | 3 | 3.5 | 4 | 4.5 | 4.75 | 5 |
| $\beta_{12}, \beta_{21}$ | 2.5 | 2.5 | 2.5 | 2.5 | 2.5 | 2.5 | 2.5 |
| $\beta_{22}$ | 2.4 | 2 | 1.5 | 1 | 0.5 | 0.25 | 0 |
| $k$ | 0.495 | 0.475 | 0.45 | 0.427 | 0.403 | 0.393 | 3.82 |

For each of these combination of parameters, we determine *x* and *y* from system (2.7), (2.8), and the corresponding values 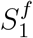 and 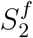. The results of these calculations are shown in Figure 1. The final value of the first susceptible sub-population slowly decreases, for the second sub-population increases. The final value of the total susceptible population increases almost 3 times between the minimal value for the homogeneous population and the maximal value reached for *β*_22_ = 0.

**Figure 1:**
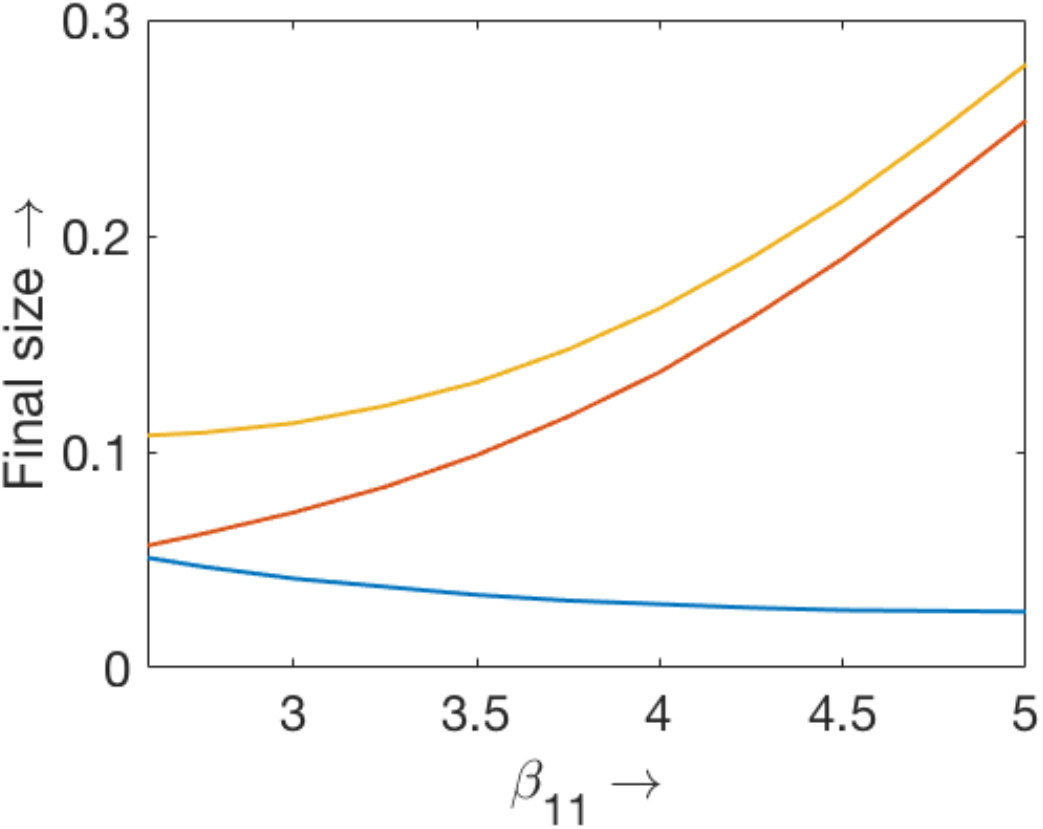
Final value of the total susceptible population and in the two sub-classes as functions of 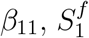 - lower curve,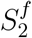 - middle curve, their sum - upper curve. The values of parameters: *β*_22_ = 5 − *β*_11_, *β*_12_ = *β*_21_ = 2.5, *σ* = 1.

In the next example, *β* = 1.5, *σ* = 1, *λ* = 0.5, *R*_0_ = 1.5. The final susceptible for the homogeneous population is *S*^*f*^ = 0.417. The maximal total susceptible for the heterogeneous population is reached for *β*_11_ = 3 and equals *S*^*f*^ = 0.587. The ratio of the maximal and minimal value decreases with the decrease of *R*_0_ but their difference remains approximately the same as in the previous example.

### 2.3 Maximum number of infected

#### Maximal number of infected individuals a for homogeneous population

If we assume that *β*_*ij*_ = *β* for all *i, j* = 1, 2 for some *β*, and *σ*_1_ = *σ*_2_, then system (2.1)-(2.4) can be reduced to the system

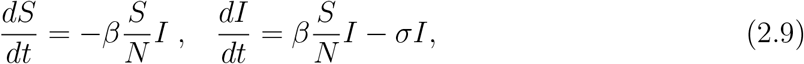

where *S* = *S*_1_ + *S*_2_, *I* = *I*_1_ + *I*_2_. Denote by *t*_*m*_ the time of maximum of *I*(*t*), and by *I*_*m*_ the maximal value, *I*_*m*_ = *I*(*t*_*m*_), and *S*_*m*_ = *S*(*t*_*m*_). Integrating the sum of the equations

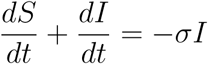

from 0 to *t*_*m*_, we get

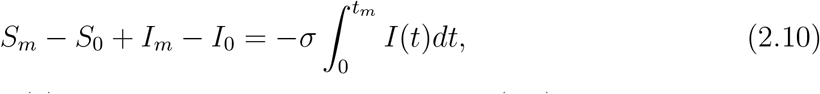

where *S*_0_ = *S*(0) = *N, I*_0_ = *I*(0). Next, from the first equation in (2.9),

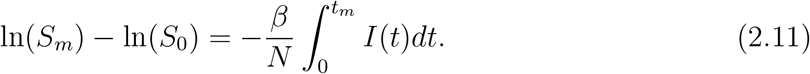

From equations (2.10), (2.11),

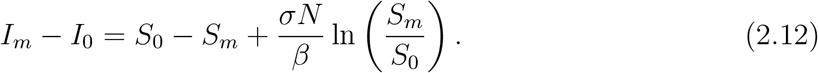

From the second equation in (2.9), since the derivative equals 0 at *t* = *t*_*m*_, we find *S*_*m*_ = *σN/β*. Using the notation *R*_0_ = *β/σ*, from (2.12),

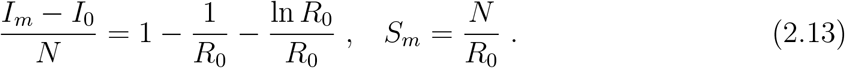

If *R*_0_ = 1, then *I*_*m*_ = *I*_0_, *S*_*m*_ = *N*. For *R*_0_ *>* 1, *I*_*m*_ *> I*_0_ and *S*_*m*_ *< N*.

#### Maximal number of infected individuals in a heterogeneous population

In order to find the maximal number of infected individuals in the heterogeneous population, we consider an approximation 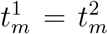, where 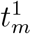 is the time of maximum of *I*_1_(*t*) and 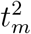 of *I*_2_(*t*). Numerical simulations show that these times to maximum are close to each other. Integrating equations (2.6) from 0 to *t*_*m*_, we obtain:

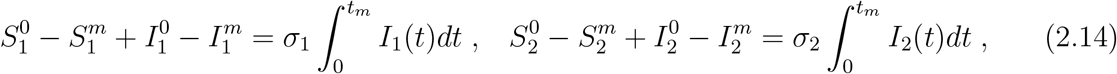

where 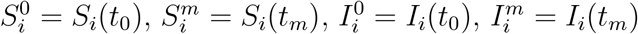, *i* = 1, 2. Next, we divide equation (2.1) by *S*_1_, equation (2.2) by *S*_2_ and integrate:

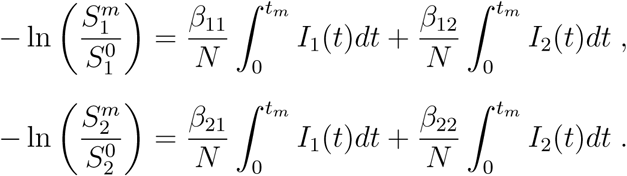

Taking into account (2.14), we obtain

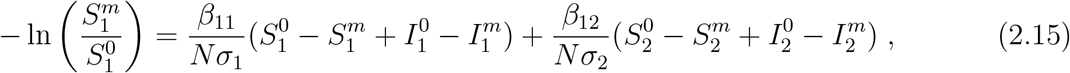

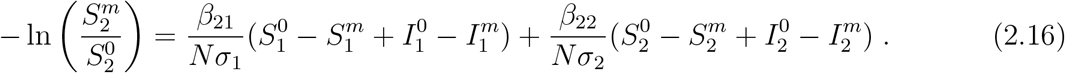

Assuming that 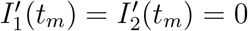, we get from (2.3), (2.4):

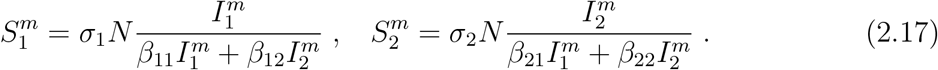

We suppose that 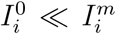, *σ*_1_ = *σ*_2_. Set 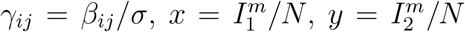. With this notation and (2.17), equations (2.15), (2.16) can be written in the following form:

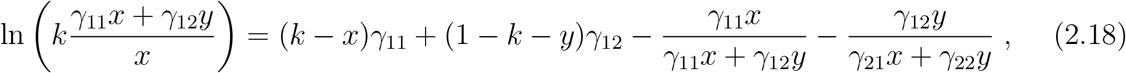

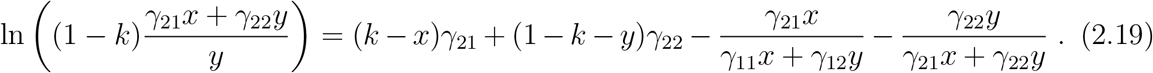

Solving this system of equations, we find *x* and *y* and, consequently, 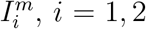. We then use formulas (2.17) to determine 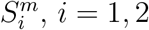.

Figure 2 shows the comparison of the values 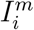 and 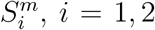 obtained from direct numerical simulations and found by the approximate analytical method presented above. This approximation is more accurate for *β*_11_ *> β*_22_ (*β*_11_ + *β*_22_ = 5) which corresponds to our main assumption that the first sub-population is smaller and spreads infection faster than the second sub-population. Let us recall that the population is homogeneous if *β*_11_ = *β*_22_. The heterogeneity of the population increases with the increase of *β*_11_. The maximal current number of infected individuals decreases with the increase of *β*_11_. This effect is especially pronounced for the second sub-population.

**Figure 2:**
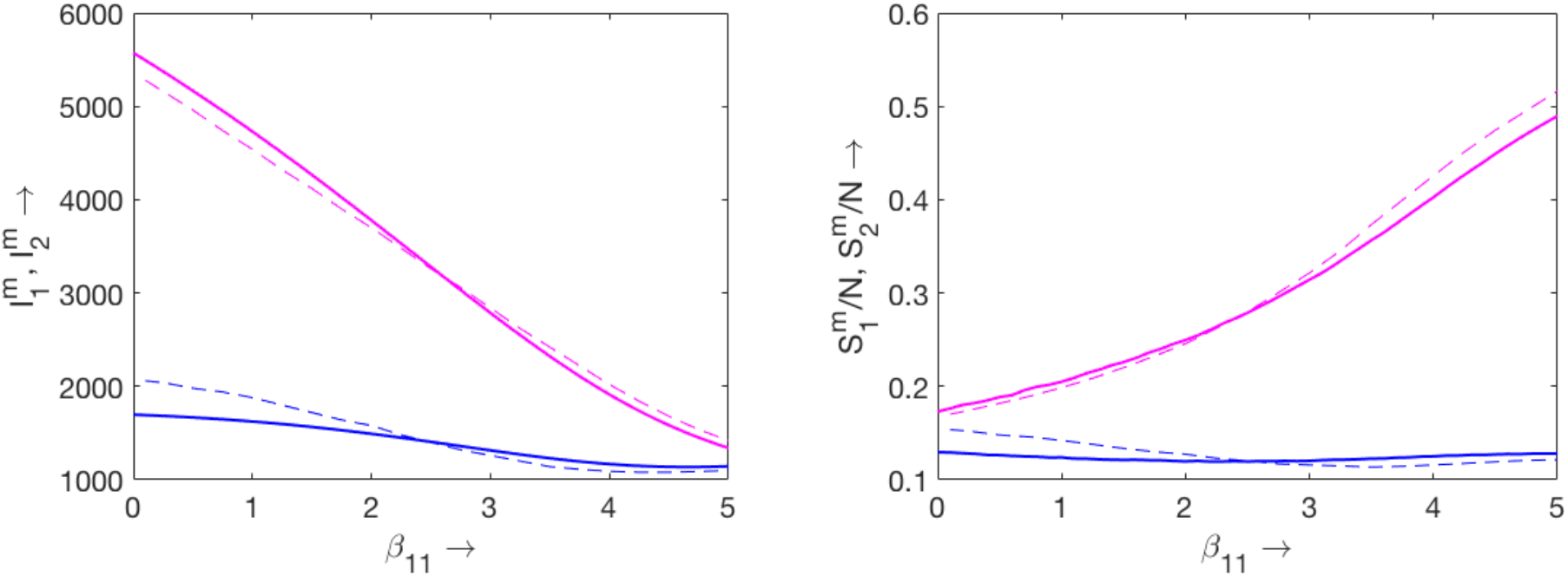
The maximal number of infected individuals (left figure) in direct numerical simulations of system (2.1)-(2.4) (solid lines) and as solution of system (2.18), (2.19) (dashed lines). The lower curves correspond to the first sub-population 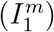 and the upper curves to the second sub-population 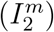. The corresponding values of the number of susceptible individuals (right figure) in numerical simulations of system (2.1)-(2.4) (solid lines) and by formulas (2.17). The lower curves correspond to the fist sub-population 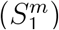 and the up-per curves to the second sub-population 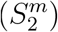. The values of parameters: *β*_22_ = 5 − *β*_11_, *β*_12_ = *β*_21_ = 2.5, *σ* = 0.1. The value of *k* is chosen in such a way that the basic reproduction number *R*_0_ = 2.5 is the same in all simulations (see the explanation in the text).

### 2.4 Collective immunity

The notion of collective immunity implies that epidemic progression slows down due to the decrease of the number of susceptible individuals. Let us give a more precise definition for the homogeneous population implying that collective immunity begins at the moment of time when the number of infected individuals reaches its maximum, *t* = *t*_*m*_, *I*_*m*_ = *I*(*t*_*m*_). The number of infected individuals begins to decrease after this time. The results of the previous section allow us to determine the exact value of *I*_*m*_ and *S*_*m*_ but not *t*_*m*_. In order to find an approximate value of *t*_*m*_ we use the approximation *I*(*t*) = *I*_0_*e*^*λt*^ within the time interval *I*_0_ *≤ I*(*t*) *≤ I*_*m*_. Then

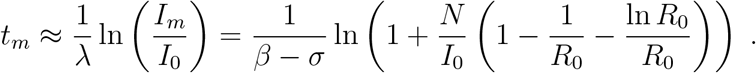

In the case of heterogeneous population, the total number of infected individuals *I*(*t*) = *I*_1_(*t*) + *I*_2_(*t*) has a single maximum at some *t* = *t*_*m*_ though the maxima of each component *I*_1_(*t*) and *I*_2_(*t*) are reach at some close but different times 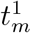 and 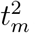, respectively. We consider that collective immunity begins at time *t* = *t*_*m*_. In the approximate analytical solution considered above we assume that 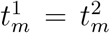 Under this approximation, the time of the beginning of collective immunity can be determined similarly to the homogeneous population but with more cumbersome calculations related to solution of system (2.18), (2.19).

### 2.5 Vaccination

The result of vaccination of a heterogeneous population essentially depends on the distribution of vaccinated individuals between different population groups. The epidemic is mainly spread by the first sub-population (HT), and their vaccination efficiently decreases the number of infected individuals.

We assume that vaccination is fully efficient in the sense that vaccinated individuals do not become infected. In order to model the action of vaccination at time *t* = *t*_0_, we set

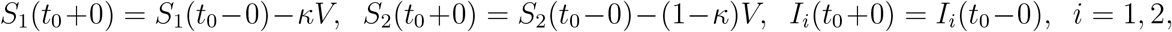

where *V* is the number of vaccinated, *κ* is part of vaccinated in the first sub-population, (1 − *κ*) in the second sub-population. System (2.1)-(2.4) is considered for *t > t*_0_ with the indicated initial conditions at *t* = *t*_0_.

Figure 3 (left) shows the total number of infected individuals *I*_*T*_ depending on vaccination. The total number of infected is calculated by the formula: 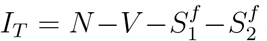 In numerical simulations we set 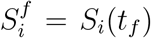, *i* = 1, 2, where *t*_*f*_ is the final time of epidemic defined as time when the number of infected individuals becomes less than 1. Let us recall, that *I*_*i*_(*t*) converge to 0 as *t*→ ∞ but these functions remain positive for any finite time. Taking into account that these variables signify the number of individuals, the epidemic can be considered as finished at *t* = *t*_*f*_ defined above.

**Figure 3:**
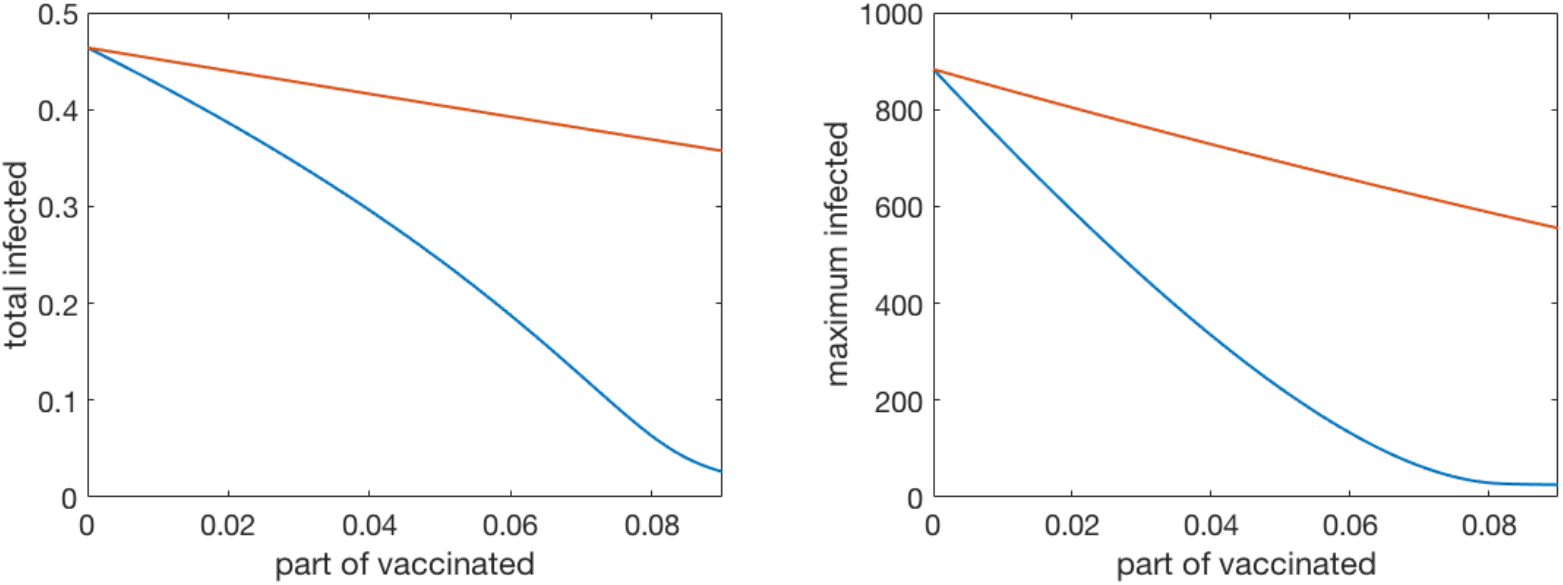
The total number of infected individuals (left figure) in numerical simulations of system (2.1)-(2.4) at the end of epidemic depending on the proportion of vaccinated individuals *V* to the total population *N*. The lower curve corresponds to the vaccination of the first sub-population (*κ* = 1) and the upper curve to the vaccination of the second sub-population (*κ* = 0) with the same total number of vaccinated individuals. The maximal current number of infected individuals (right figure) depending on the proportion of vaccinated individuals *V* to the total population *N*. The lower curve corresponds to the vaccination of the first sub-population (*κ* = 1) and the upper curve to the vaccination of the second sub-population (*κ* = 0) with the same total number of vaccinated individuals. The values of parameters: *β*_11_ = 4, *β*_22_ = 1, *β*_12_ = *β*_21_ = 2.5, *σ* = 0.1, *k* = 0.1, *t*_0_ = 5.

We compare two cases, where all vaccinated belong to the first sub-population (*κ* = 1) or all of them belong to the second sub-population (*κ* = 0). In the first case, the influence of vaccination on the total number of infected individuals at the end of epidemic is essentially stronger than in the second case. If we take as example *V* equal 5% of the total population (*V/N* = 0.05), then *I*_*T*_ reduces from 0.45*N* (without vaccination) to 0.25*N*, that is almost twice. At the same time, if only the second population is vaccinated, then the reduction is only 5%, that is the same as the number of vaccinated.

This difference becomes even more essential for the maximal current number of infected individuals (Figure 3, right). More detailed data on the results of vaccination are presented in Appendix. These results depend on the values of parameters, in particular on the parameter *k* characterizing the proportion between the two sub-populations. However, the general tendency is the same as presented above: vaccination of the first sub-population is much more efficient. It is also interesting to note that vaccination increases the final time of epidemic.

## 3 Full model

### 3.1 Model of heterogeneous population

We consider conventional compartmental approach to model the epidemic progression with the following classes of population: susceptible individuals *S*, exposed (with viral load) but not yet infectious *E*_1_, exposed infectious (no yet symptoms) *E*_2_, infected symptomatic *I*_*s*_, infected asymptomatic *I*_*a*_, quarantined *Q*, hospitalized *J*, recovered *R*. Exposed infectious and infected asymptomatic individuals are in some sense similar to each other because they are infectious but do not manifest symptoms. However, the rate of disease transmission and the duration of these stages for them can be different.

The susceptible population can be heterogeneous with respect to a variety of characteristics: age classes, their activity including education, professional, retired, medical workers [18, 19, 20]. We will study how the heterogeneity of the population can influence the final size and duration of epidemic. For simplicity of presentation and analysis we restrict ourselves to two subclasses of susceptible individuals *S*_1_ and *S*_2_. According to this separation on subclasses, we introduce the corresponding subclasses in the groups *E*_1_, *E*_2_, *I*_*s*_, and *I*_*a*_, while *Q, J*, and *R* remain homogeneous. Under these assumptions, we obtain the following equations for *S*_1_ and *S*_2_:

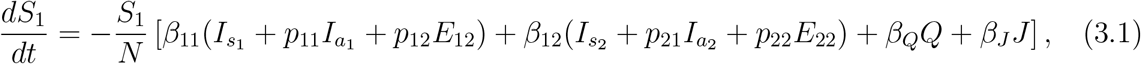

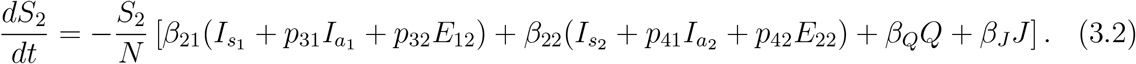

Here 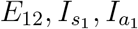 and 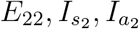 are the subclasses of the corresponding classes *E*_2_, *I*_*s*_, *I*_*a*_; *β*_*ij*_ are the coefficients characterizing the intensity of infection transmission between the classes *S*_*i*_ and 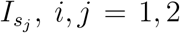; the coefficients *p*_*ij*_, *i* = 1, 2, 3, 4, *j* = 1, 2 show how the coefficients of infection propagation change for the classes 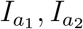 and *E*_21_, *E*_22_ in comparison with 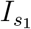 and 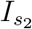. Finally, the coefficients *β*_*Q*_ and *β*_*J*_ characterize infection progression due to the interaction with the classes *Q* and *J*.

The corresponding equations for the classes *E*_11_, *E*_21_ have the following form:

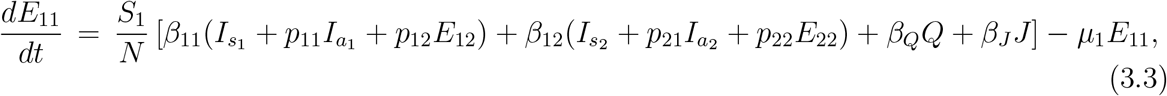

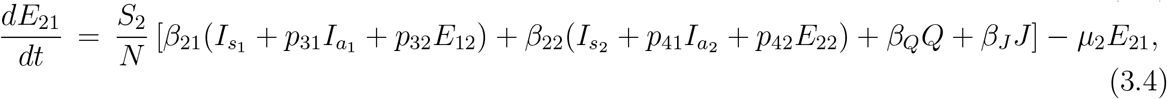

where *µ*_1_ and *µ*_2_ are the rates at which *E*_11_ and *E*_21_ progress to the infectious exposed compartments *E*_12_ and *E*_22_, respectively. Next,

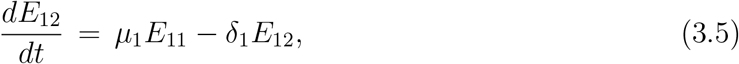

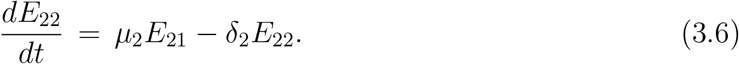

The coefficients *δ*_1_ and *δ*_2_ characterize the transition from exposed to infected classes. The equations for the infected classes are as follows:

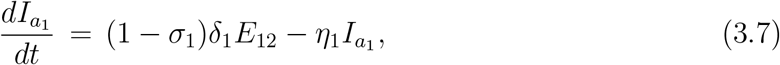

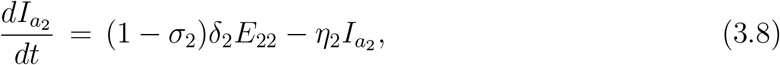

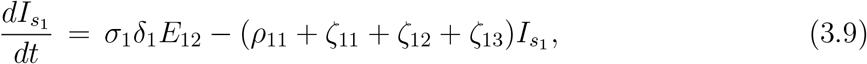

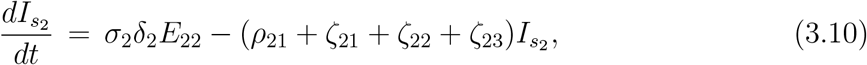

where *σ*_1_ and *σ*_2_ determine the the proportions between the classes of symptomatic and asymptomatic individuals, 0 *< σ*_*i*_ *<* 1, *i* = 1, 2.

Symptomatic infected individuals can become quarantined, hospitalized or recover, and asymptomatic infected recover without hospitalization. Symptomatic individuals move to the quarantine class, and the rate of their transfer to hospital and recovered class are ξ_1_ and ξ_2_, respectively. The equation for the quarantined class writes:

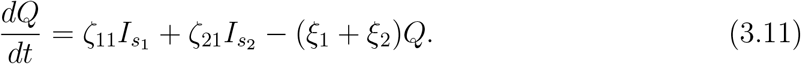

Hospitalized individuals can recover or die with the rates *ν* and *ρ*_2_:

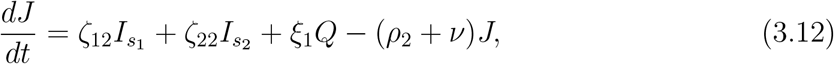

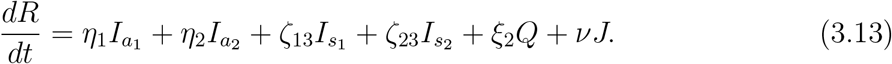

This model was introduced in [9] where the basic reproduction number and the final size of epidemic were found. It was also used to fit the data on the Covid-19 epidemic in some countries before and during the lockdown and to determine the parameters by fitting the numerical simulation with the epidemiological data. Furthermore, most sensitive parameters were estimated. We will use this model and parameter values in the next section in order to study the influence of vaccination on epidemic progression, as illustrative example.

### 3.2 Characterization of epidemic progression

In this section we present the results of numerical simulations of system (3.1)-(3.13). Figure 5 shows the evolution in time of the two susceptible populations *S*_1_(*t*) (left figure) and *S*_2_(*t*) (right figure) for different values of parameter *k* = *N*_1_*/N*. Larger values of *k* corresponds to the increase of the proportion of the first sub-population for which the disease transmission rate is more intensive. We observe from the figure that increasing *k* leads to the decrease of the final time of epidemic. The final size of the first sub-population remains approximately constant. This is due to the fact that epidemic is basically transmitted by the first sub-population, and it is finished when this sub-population reaches collective immunity. The final size of the second sub-population decreases with the increase of *k* (Figure 6, left) but it remains above the level of collective immunity. These final sizes can be found from the analytical formulas obtained in [9].

**Figure 4:**
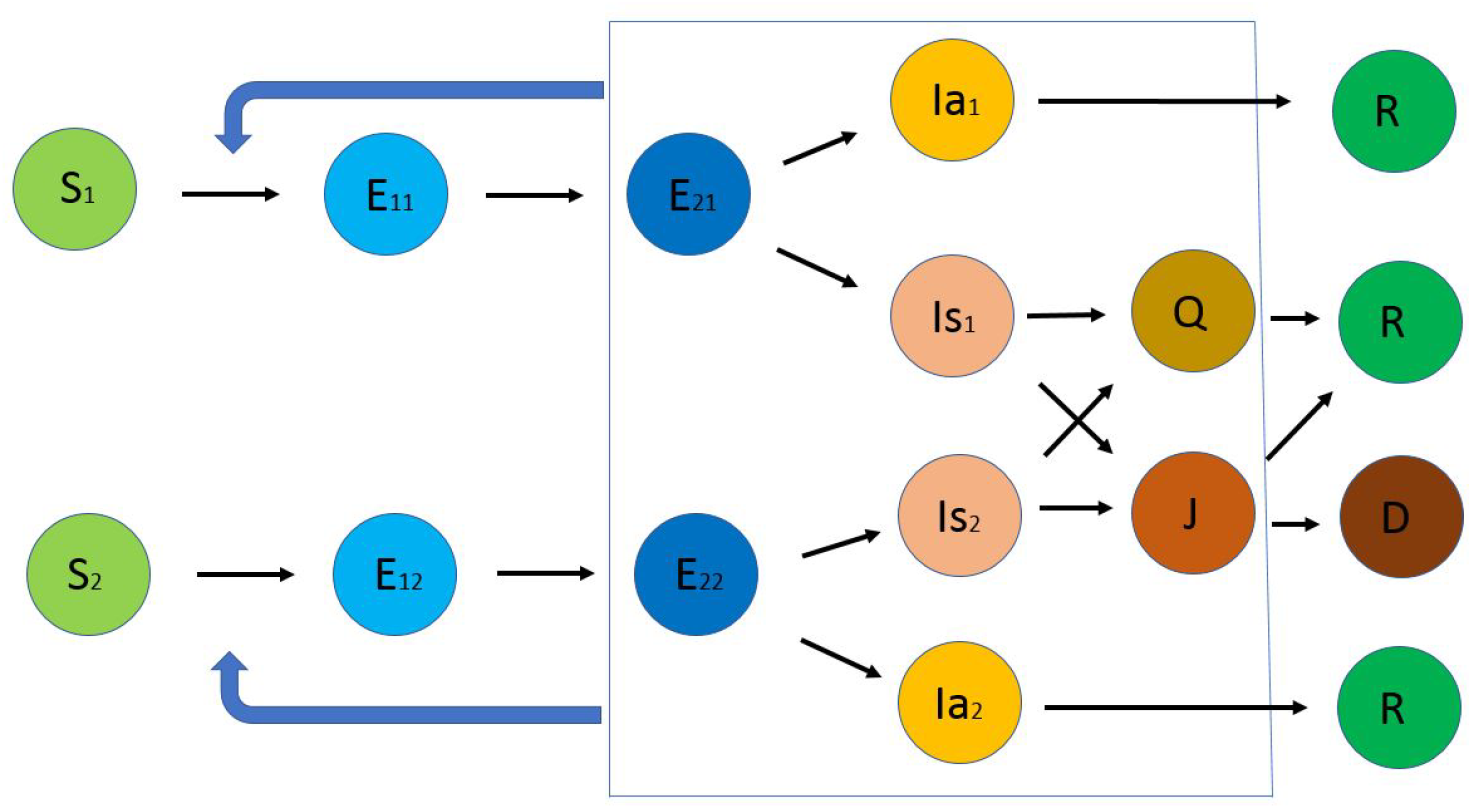
Schematic representation of the model with different classes of individuals. Two subclasses of susceptible give respectively exposed non-infectious, exposed infectious, infected symptomatic and asymptomatic. There are unique classes of quarantined, hospitalized, recovered, and dead.

**Figure 5:**
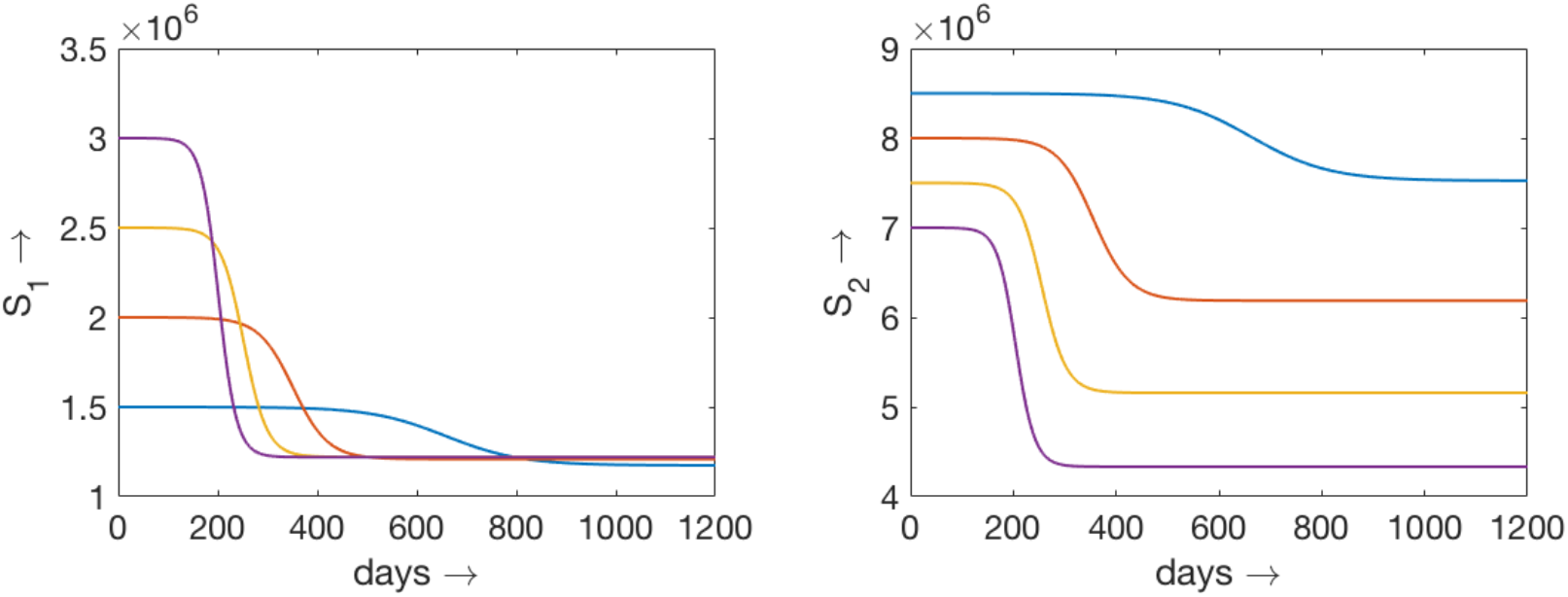
Numerical simulations of system (3.1)-(3.13). The evolution of the sub-populations *S*_1_(*t*) (left) and *S*_2_(*t*) in time for different values of *k*. The values of the coefficients *β*_*ij*_ are as follows: *β*_11_ = 4, *β*_22_ = 1, *β*_12_ = *β*_21_ = 2.5. The values of other parameters are given in the appendix.

**Figure 6:**
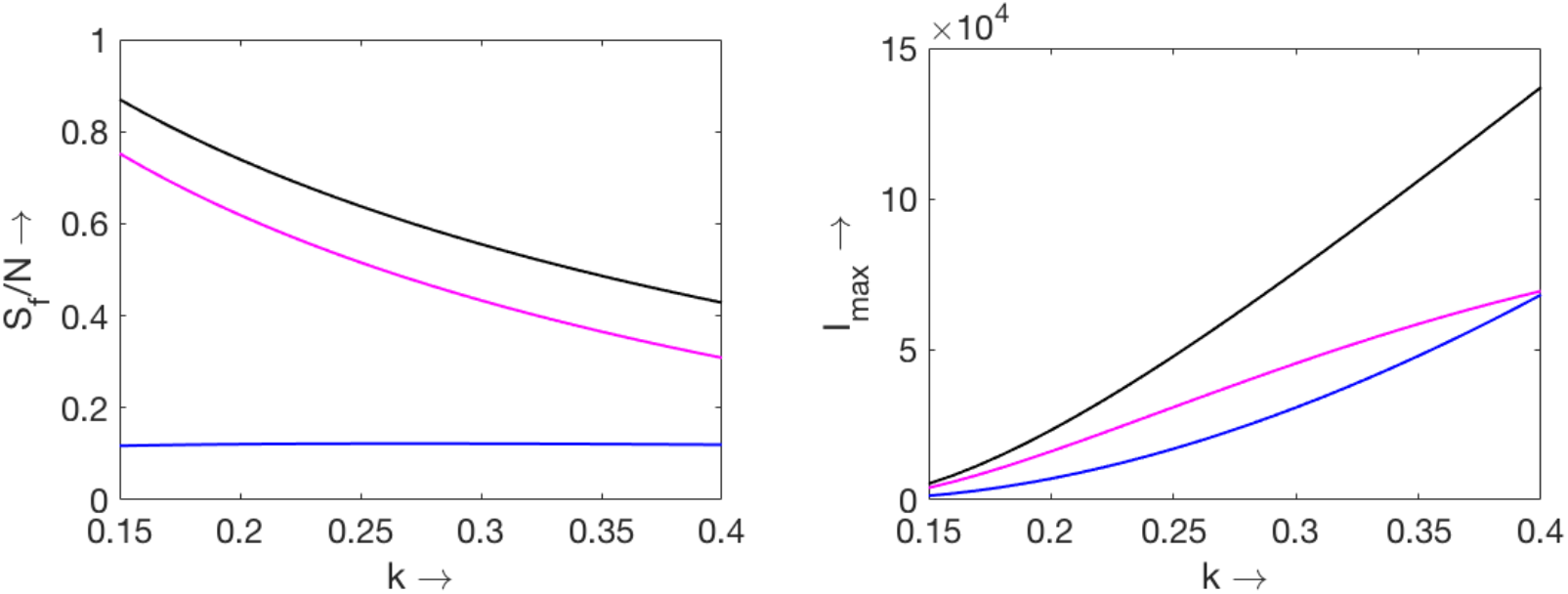
The final size of susceptible classes for different values of *k* (left). The upper curve shows the total number of susceptible, the middle curve corresponds to *S*_2_(*t*) and the lower curve to *S*_1_(*t*). The maximal number of infected individuals for different values of *k* (right). The upper curve shows the total number of infected (symptomatic plus asymptomatic), the middle curve corresponds to the second sub-population and the lower curve to first sub-population. The values of the coefficients *β*_*ij*_ are as follows: *β*_11_ = 4, *β*_22_ = 1, *β*_12_ = *β*_21_ = 2.5. The values of other parameters are given in the appendix.

The maximal values of infected individuals increase with the increase of *k* (Figure 6, right). We consider here the sum of symptomatic and asymptomatic classes, 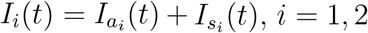. Though the first sub-population *N*_1_ = *kN* is essentially less than the second one, *N*_2_ = (1 − *k*)*N* for small *k*, the maximal values of infected individuals, 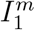 and 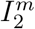 are close to each other because disease transmission occurs faster in the first sub-population.

## 4 Application to the Covid-19 epidemic

### 4.1 Coefficient of social interaction

The heterogeneity of the population with respect to the disease transmission is related to multiple factors. Among them different age, professional and social groups, various religious and cultural traditions which can influence people behavior with respect to the measures of social distancing and vaccination. Detailed description of all these different groups would essentially complicate the model and would increase the number of parameters difficult to estimate. Therefore, we propose to consider only two cumulative groups. One of them includes people with high disease transmitting potential (HT) and another one with low disease transmitting potential (LT). According to the models we consider here, these two classes are represented by *S*_1_ and *S*_2_ respectively. They differ by the values of parameter *β*_*ij*_ in the expression *β*_*ij*_*I*_*i*_*S*_*j*_*/N, i, j* = 1, 2. These are effective parameters characterizing the frequency of contacts between infected and susceptible individuals and the rate of infection transmission. For the two groups *N*_1_ (HT) and *N*_2_ (LT) of the whole population *N*, there are the corresponding subclasses of susceptible *S*_1_ and *S*_2_, exposed and infected. Hence, there are four different coefficients: *β*_11_ characterizes the interaction inside HT, *β*_22_ inside LT, and *β*_12_, *β*_21_ between the groups.

We consider the parameter *k* = *N*_1_*/N* already used in the previous sections. For *k* = 0, there is a single group LT, for *k* = 1 another single group HT. We will consider the values of *k* between 0 and 1. This parameter characterizes the distribution of total population into two groups and influences the intensity of social interactions.

We determine the values of parameters *β*_*ij*_ and *k* from the Covid-19 data. In the beginning of the epidemic, before lockdown, there were no measures of social distancing. We suppose that the whole population belonged to the first group (HT). We neglect here the heterogeneity of the population with respect to the frequency of contacts. Fitting the data on the epidemic progression allows us to determine the coefficient *β*_11_. Its value can be different in different countries. Next, we suppose that during the first lockdown the whole population respected the measures of social distancing and belonged to the second group (LT). As before, fitting the data allows us to determine *β*_22_. In the data fitting we used the model presented in Section 3 [9]. The values of the other parameters were determined from the available data.

After the first lockdown, the measures of social distancing were partially preserved. These restrictions differed between the countries and evolved in time. They were less strict than during the first lockdown allowing the emergence of two cumulative groups *N*_1_ and *N*_2_ described above. Simplifying this characterization of the population, we suppose that the first group (HT) is similar to the population before lockdown, and it is characterized by the coefficient *β*_11_ described above. The second group (LT) is similar to the population during the lockdown, and it is characterized by the coefficient *β*_22_. We set the values of the coefficients *β*_12_ and *β*_21_ characterizing the interactions between the groups according to the formula *β*_12_ = *β*_21_ = (*β*_11_ + *β*_22_)*/*2. It is an empiric relation which cannot be determined from the data. We will discuss it below.

Next, we determine the value of the coefficient *k* fitting the data after lockdown. Figure 7 shows consecutive stages of epidemic progression in Germany with the first stage (before lockdown), second stage (during lockdown), third stage (June-July, 2020), and the fourth stage (September-November, 2020). According to the method described above, we get *β*_11_ = 3.95, *β*_22_ = 1.05, *β*_12_ = *β*_21_ = 2.5. Fitting the data in June-July, we find *k* = 0.1, that is the first group (HT) represents 10% of the whole population. We then continued the simulation for the period September-November with two different values of *k*: the same as before, *k* = 0.1 (left figure), and *k* = 0.2 (right figure). Increase of *k* shows a rapid growth of the number of infected. Let us note that this simulation was done in July, 2020 [9], and it gave a reasonably good prediction of the epidemic progression during the fourth stage. Increase of the coefficient of social interaction *k* during the fourth stage is related to the beginning of the academic year and the intensification of professional activity after summer vacation.

**Figure 7:**
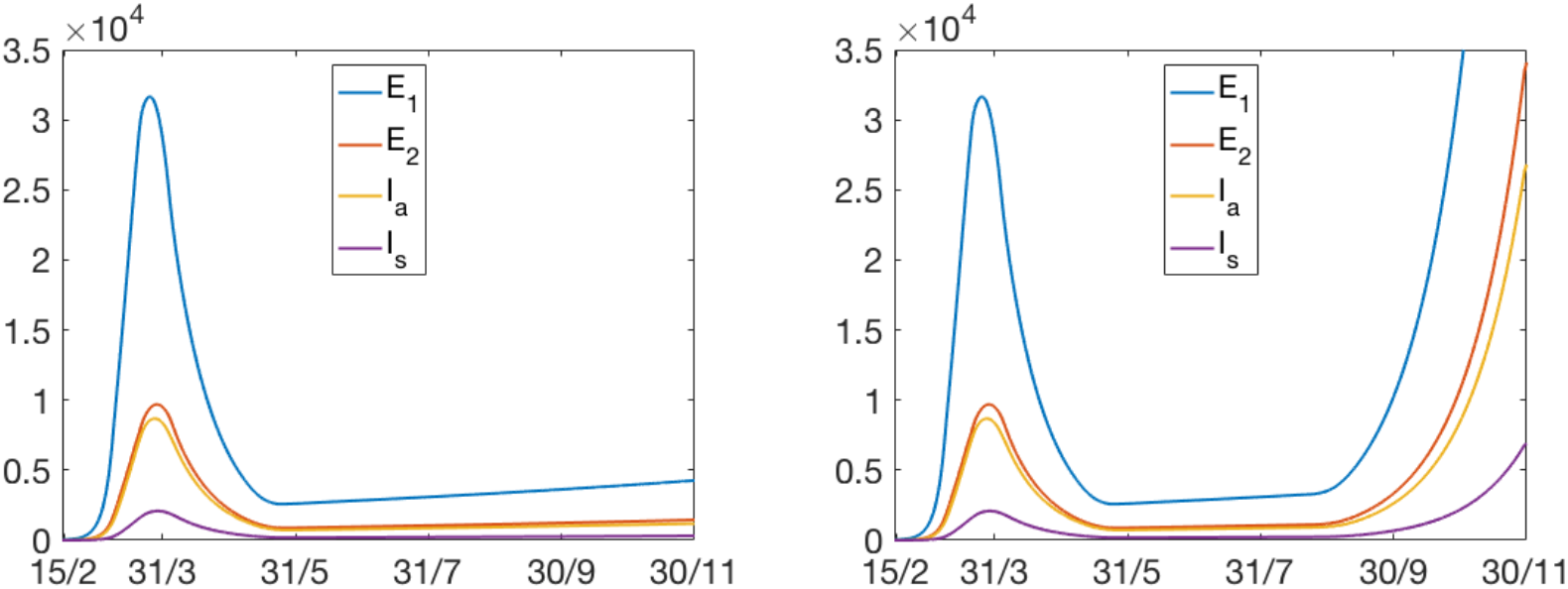
Numerical simulations of epidemic progression in Germany with system (3.1)-(3.13). The values of the coefficients *β*_*ij*_ are as follows: *β*_11_ = 3.95, *β*_22_ = 1.05, *β*_12_ = *β*_21_ = 2.5 (see the explanation in the text). The values of other parameters are given in the appendix. The values of *k*: 1 in February-March 2020 (before lockdown), 0 in April-May (lockdown), 0.1 in June-August (after lockdown), 0.1 in September-November (left) and 0.2 in September-November (right). Reprinted from [9] with permission.

Another example is shown in Figure 8. Fitting of data for Israel gives *β*_11_ = 2.91, *β*_22_ = 0.3, *β*_12_ = *β*_21_ = 1.17. The value of *k* after lockdown varied from 0.1 to 0.3. The estimates of the coefficients *β*_*ij*_ for some European countries are presented in [9]. The values around *β*_11_ = 4 and *β*_22_ = 1 are quite specific, and we used them in the previous sections of this work. The value of *k* after the first lockdown usually changes between 0.1 and 0.3. We will use these characteristic values of parameters in the next subsection in order to study the influence of vaccination on the epidemic progression.

**Figure 8:**
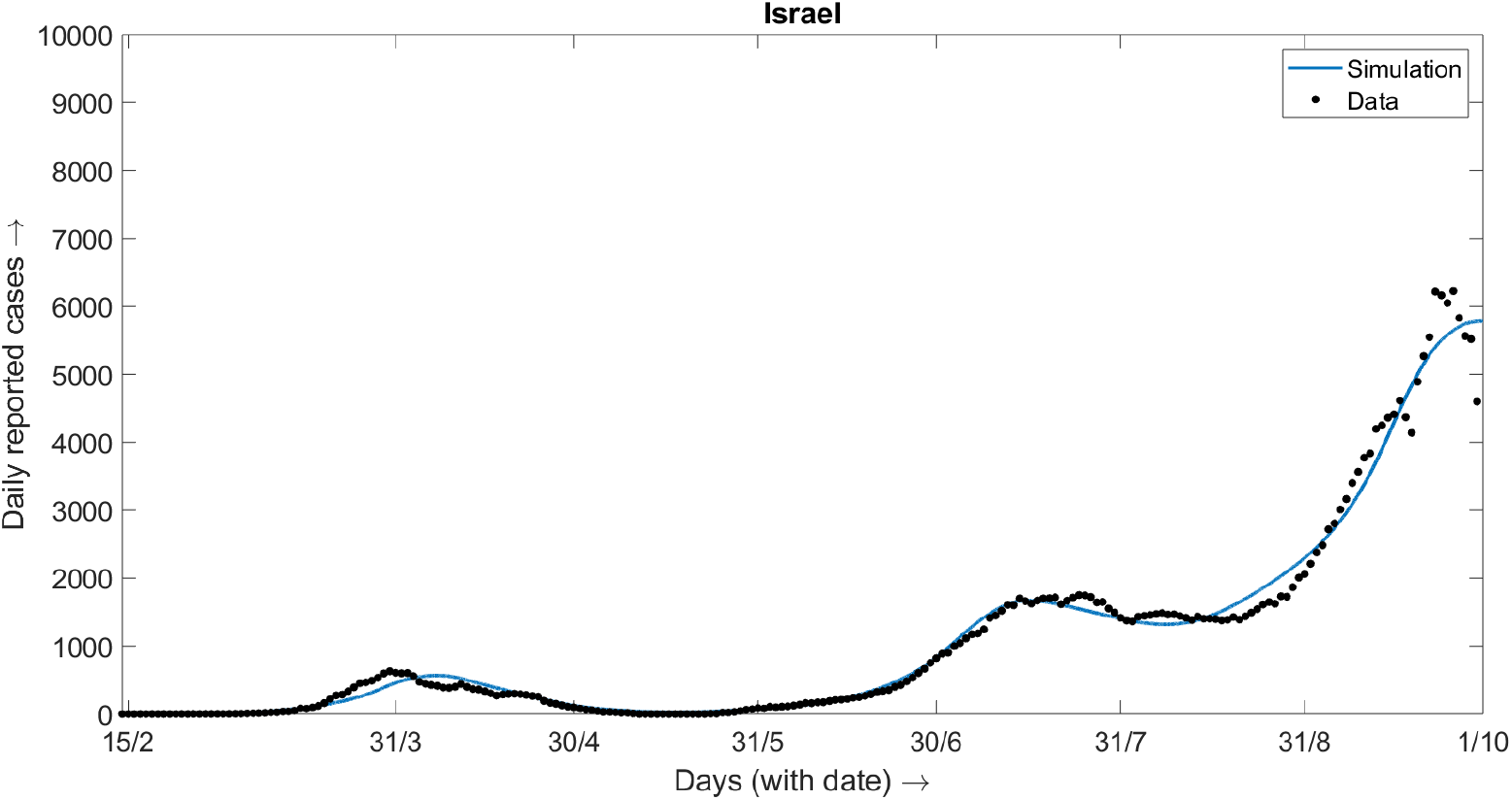
Numerical simulations of epidemic progression in Israel with system (3.1)-(3.13). The values of the coefficients *β*_*ij*_ are as follows: *β*_11_ = 2.91, *β*_22_ = 0.3, *β*_12_ = *β*_21_ = 1.17. The values of other parameters are given in the appendix. The values of *k* after lockdown changes from 0.1 to 0.3.

### 4.2 Vaccination

We proceed to the effect of vaccination on the epidemic progression. Similar to the modelling approach considered in Section 2, we apply vaccination at some time *t* = *t*_0_ and model it by decreasing the number of susceptible individuals:

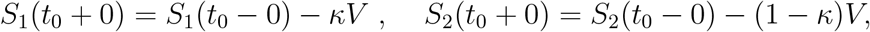

while all other classes do not change. Here *κ* is the proportion of all vaccinated individuals *V* in the first sub-population and (1 − *κ*) in the second one.

Figures 9 and 10 show the influence of the number of vaccinated individuals and of their distribution among the two sub-population on the total and maximum number of infected individuals. The total number of infected individuals in the first sub-population is determined as 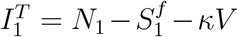 and in the second sub-population 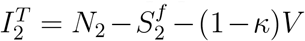, where 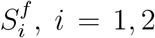, *i* = 1, 2 are the final numbers of susceptible individuals, 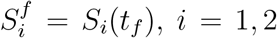, *i* = 1, 2 where *t*_*f*_ is time at which 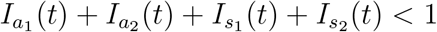. The maximal current number of infected individuals in each sub-class are defined as the maximum of the function 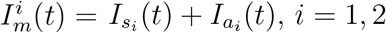.

**Figure 9:**
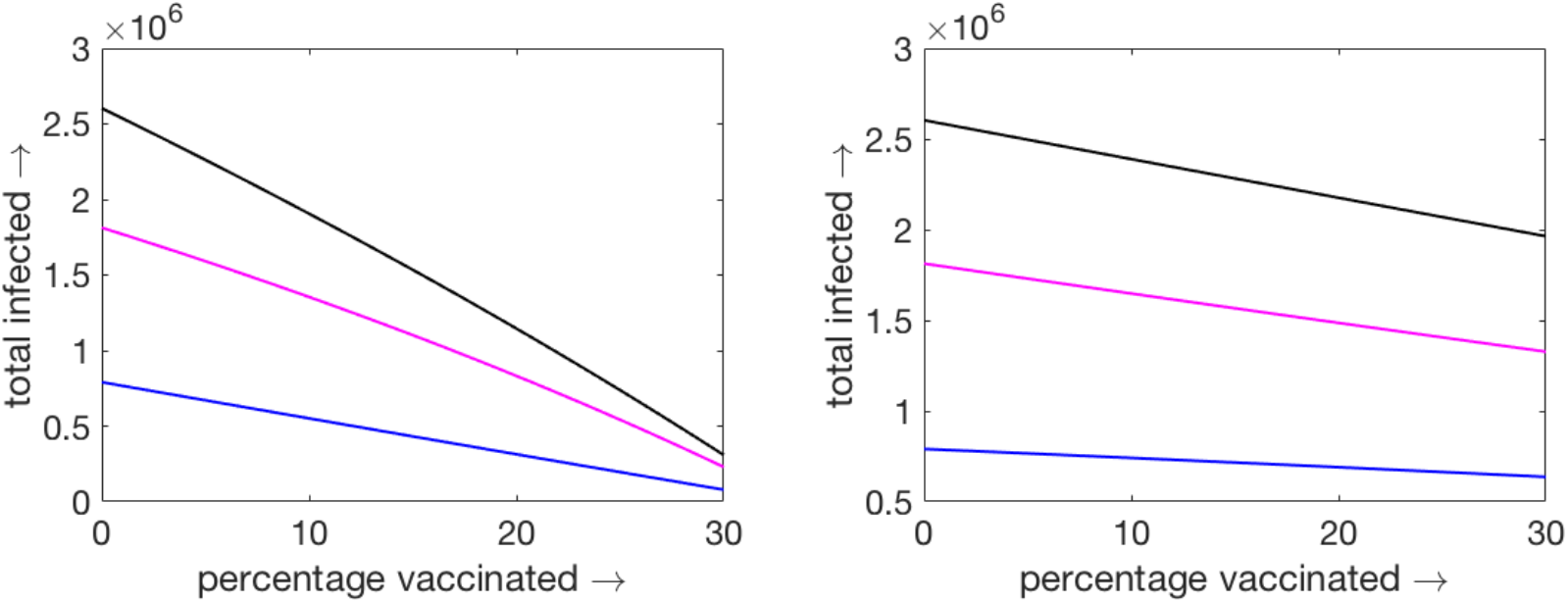
Total number of infected individuals at the end of epidemic as a function of the number vaccinated individuals applied only to the first sub-population (*κ* = 1, left figure) or only to the second sub-population (*κ* = 0, right figure). The percentage of vaccinated individuals is counted with respect to *N*_1_ in both cases. The lower curve shows the total number on infected individuals in the first sub-population, the middle curve in the second sub-population, and the upper curve their sum.

**Figure 10:**
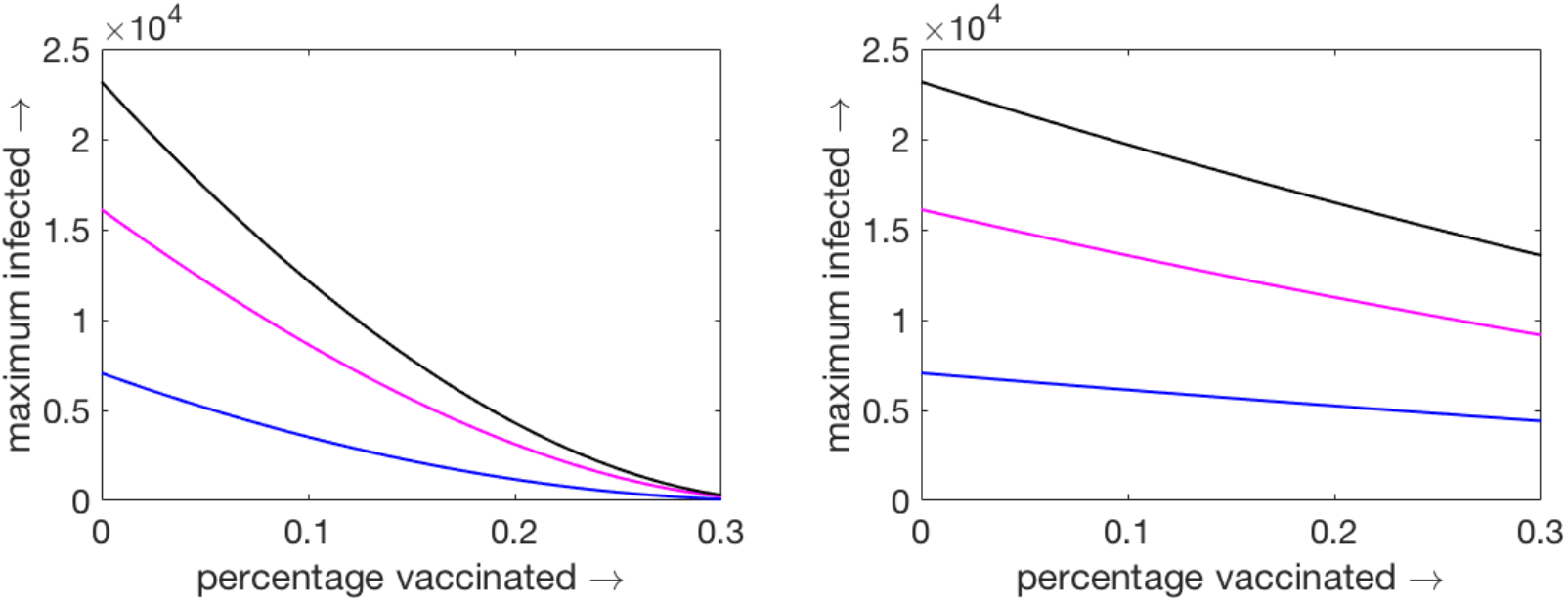
Maximal current number of infected individuals as a function of the number vaccinated individuals applied only to the first sub-population (*κ* = 1, left figure) or only to the second sub-population (*κ* = 0, right figure). The percentage of vaccinated individuals is counted with respect to *N*_1_ in both cases. The lower curve shows the maximal number on infected individuals in the first sub-population, the middle curve in the second sub-population, and the upper curve their sum.

The result of the vaccination strongly depends on its distribution between the two sub-populations. If vaccination is applied to the first sub-population (*κ* = 1), it decreases the total number of infected individuals much stronger than if it is applied to the second sub-population (*κ* = 0) (Figure 9). In this example, *k* = 0.2 and *N*_1_ = 0.2*N*. The percentage of vaccinated individuals is measured with respect to *N*_1_. So, 30% of vaccination in HT class correspond to 6% of the total population *N*. In this case, the total number of infected individuals at the end of epidemic decreases 6 times if the vaccination is applied to the first sub-population, compared to the same number of vaccination (6% of total population) is applied to the second sub-population. This striking difference shows that in the first case vaccination acts to stop epidemic progression while in the second case it only protects vaccinated individuals from infection. This difference is even more essential for the current maximal number of infected individuals (Figure 10).

## 5 Discussion

### Actual stage and epidemic waves

At the end of the first year of the Covid-19 epidemic, some of its properties are already sufficiently well understood. Among them, it is now clear that the epidemic progression obeys the usual epidemiological laws, and it can be described by conventional epidemiological models. There is a large body of research devoted to the description of the first stage of epidemic with such models, to the determination of the basic reproduction number and to related questions, basically with ODE models (see [16] and the references therein) but also with individual based models [17].

Another important observation is that, due to relatively high mortality rate and proportion of severe cases, the epidemic cannot be left to follow its natural route to collective immunity and extinction. National health systems become rapidly saturated and fail to treat not only coronavirus patients but the whole population. Therefore, the only available method up to now consists in the introduction of the measures of social distancing, confinement, wearing mask and even strict lockdown [21].

On the other hand, these measures impose a heavy burden on the economy, and they are relaxed as soon as the epidemiological situation improves. Some time later, the epidemic progression restarts, and it becomes necessary to introduce these measures again. We now observe the second wave of epidemic spread at different countries, and they will certainly continue unless the vaccination will stop them. These oscillations in the epidemic progression can be described by the simple delay differential equation

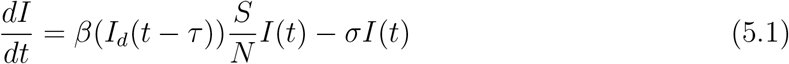

for the number of infected individuals *I* assuming that the number of susceptible individuals *S* is constant in the beginning of epidemic and *β*(*I*_*d*_) is a decreasing function of the number of new daily cases *I*_*d*_ taken with some time delay. This function described the measures of social distancing depending on the epidemiological situation. Since the number of new daily cases is usually taken proportional to the product *SI*, we obtain a closed equation with respect to *I*(*t*). During further epidemic progression, when *S* cannot be considered as constant, the combination of equation (5.1) with the other equations of the epidemiological models will describe the interaction of this oscillatory dynamics with collective immunity.

### Further epidemic progression depends on the structure of the population

At the actual stage of epidemic progression, serological tests in some countries show that there are of the order of 10% of population with antibodies [22, 23]. We are yet far from the collective immunity but we need already to take into account the variation of *S*. Moreover, vaccination expected during the next year will also change the number of susceptible and, consequently, will influence the pattern of epidemic progression.

In the beginning of epidemic, observed exponential growth of the number of infected individuals can be described for any population structure [5, 10, 24, 25]. The heterogeneity of the population becomes important at the later stages of the epidemic development when it deviates from the exponential growth and approaches the stage of collective immunity and when it decays approaching the final time, defined as time when the number of infected individuals becomes less than 1. Essentially, it indicates that no one remain in the system who can spread the epidemic.

We study the influence of the heterogeneity of the population with the model problem in Section 2. This relatively simple model allows us to determine the final size of epidemic, the total and the maximal current number of infected individuals. The latter is particularly important for the estimation of available hospitals beds. The main conclusion here is that the data on the initial epidemic stage are now sufficient to predict its further progression. In the case of the homogeneous population, conventional SIR model allows the determination of the final size of epidemic and of the maximal current number of infected individuals solely on the basis of the basic reproduction number, that is, on the basis of the initial growth rate. However, this is not the case for the heterogeneous population any more [15]. With the same initial growth rate, the final size of epidemic and the maximal current number of infected individuals strongly depend on the structure of the heterogeneous population. Moreover, the total and maximal numbers of infected individuals can change several times for realistic values of parameters determining the distribution of population into two groups.

### How to estimate the structure of the heterogeneous population?

Thus, we come to the question about the estimation of the structure of a heterogeneous population. In the context of COVID-19, it is needless to mention that the number of reasonable grouping seems to be greater than two but for the simplicity of mathematical modelling we restricted oursleves to HT and LT classes only. We use here the data on the Covid-19 epidemic for different countries. The main idea of our approach is to present the population as a combination of two groups, with high and low disease transmission potentials. The first group is related to the period before lockdown without measures of social distancing and some other control measures, and the second group to the period during lockdown when these measures were strict. In a simplified representation, these two groups can be identified by two factors: a) the number of interactions with other individuals, b) respect of the measures of social distancing (masks, sanitizers, and so on). For example, people who have their normal (as before lockdown) average interaction belong to the HT group, those who have reduced interaction (as during lockdown) belong to the LT group. Certainly, this is a simplified representation of the population because each group is heterogeneous itself, and some individuals can change their groups in different time periods. Furthermore, we approximate a gradual distribution of interactions by a binary function. However, these simplifications allow us to obtain tractable analytical results and give a simple description of the population characterized by a single parameter *k* defined as a proportion of the HT group to the whole population.

The population consisting of two groups is characterized by four parameters *β*_*ij*_, *i, j* = 1, 2 describing the intensity of disease transmission in the groups *S*_*i*_*I*_*j*_ *i, j* = 1, 2. The coefficient *β*_11_ for the disease transmission inside the HT group is obtained by fitting the data before lockdown. The coefficient *β*_22_ for the disease transmission inside the LT group is obtained by fitting the data during lockdown. However, the cross-group coefficients *β*_12_ and *β*_21_ cannot be determined from the data. We impose the assumption that *β*_12_ = *β*_21_ = (*β*_11_ + *β*_22_)*/*2. The justification of this assumption is based on the physical example of two groups of balls moving with different speeds, *v*_1_ and *v*_2_. The number of their collisions is proportional to the average speed (*v*_1_ + *v*_2_)*/*2. We are aware that this representation of the population is too simplified, and further analysis of these coefficients is needed.

Let us note that in the model problem (2.1)–(2.4), only parameters *β_ij_* are unknown, while *σ_i_* can be estimated from the data on disease duration. Therefore, assuming that the population is homogeneous before lockdown, that is all *β_ij_* are equal to each other, we have only one parameter *β*_11_ to determine by fitting the data. Similarly, a single parameter *β*_22_ should be determined from the data during the lockdown, and the single parameter *k* from the data after lockdown. A similar situation occurs for a more complete model considered in Sections 3 and 4. If we consider more detailed models with different sub-classes inside HT and LT groups and the corresponding contact matrices, then there are more parameters *β_ij_*, and they cannot be uniquely determined from the data.

### The influence of heterogeneity on vaccination

Knowing the structure of the population, we can investigate how its heterogeneity influences the results of vaccination. Vaccination is modeled as a decrease of the number of susceptible individuals. We assume here that vaccinated individuals cannot become infected, that is, that vaccination is fully efficient. The results of the vaccination strongly differ depending on whether it is applied to the HT group or to the LT group. In the first case, a relatively small part of vaccinated individuals (5 % of the total population) can reduced several times the total number of infected individuals at the end of epidemic, and even more, the maximal current number of infected individuals. Therefore, vaccination of the HT group strongly contributes to stop the epidemic in case of limited number of available doses of vaccine. Vaccination of the second group has much weaker influence on the epidemic progression. It is basically reduced to the protection of vaccinated individuals from infection. These results can be qualitatively expected but we need to determine first the structure of the population in order to give quantitative analysis of the results of vaccination. Future directions of this work will include vaccination taking into account age structure and comorbidities in order to reduce the death toll.

Let us also note that vaccination increases the final time of epidemic. In the case of vaccination of the HT group with 6 % of vaccinated individuals of the total population and *k* = 0.2, the final time of epidemic increases almost 6 times in comparison with the case without vaccination. More complete results of vaccination are presented in the appendix.

## Data Availability

The data is available. There is no dataset material online

## Acknowledgements

The first author has been supported by the RUDN University Strategic Academic Leadership Program.

## 6 Appendix

The tables below show the results of numerical simulations of the results of vaccination (full model) in three cases: vaccination is applied only to the first sub-population (HT), only to the second sub-population (LT), and in proportion 20 % HT and 80 % LT. In all cases the number of vaccinated individuals is the same. It is given in percentage of *N*_1_. The case *k* = 0.2 and 10 % of *N*_1_, for example, corresponds to 2% of the total population *N*. In all cases the vaccine is administrated on day 5.

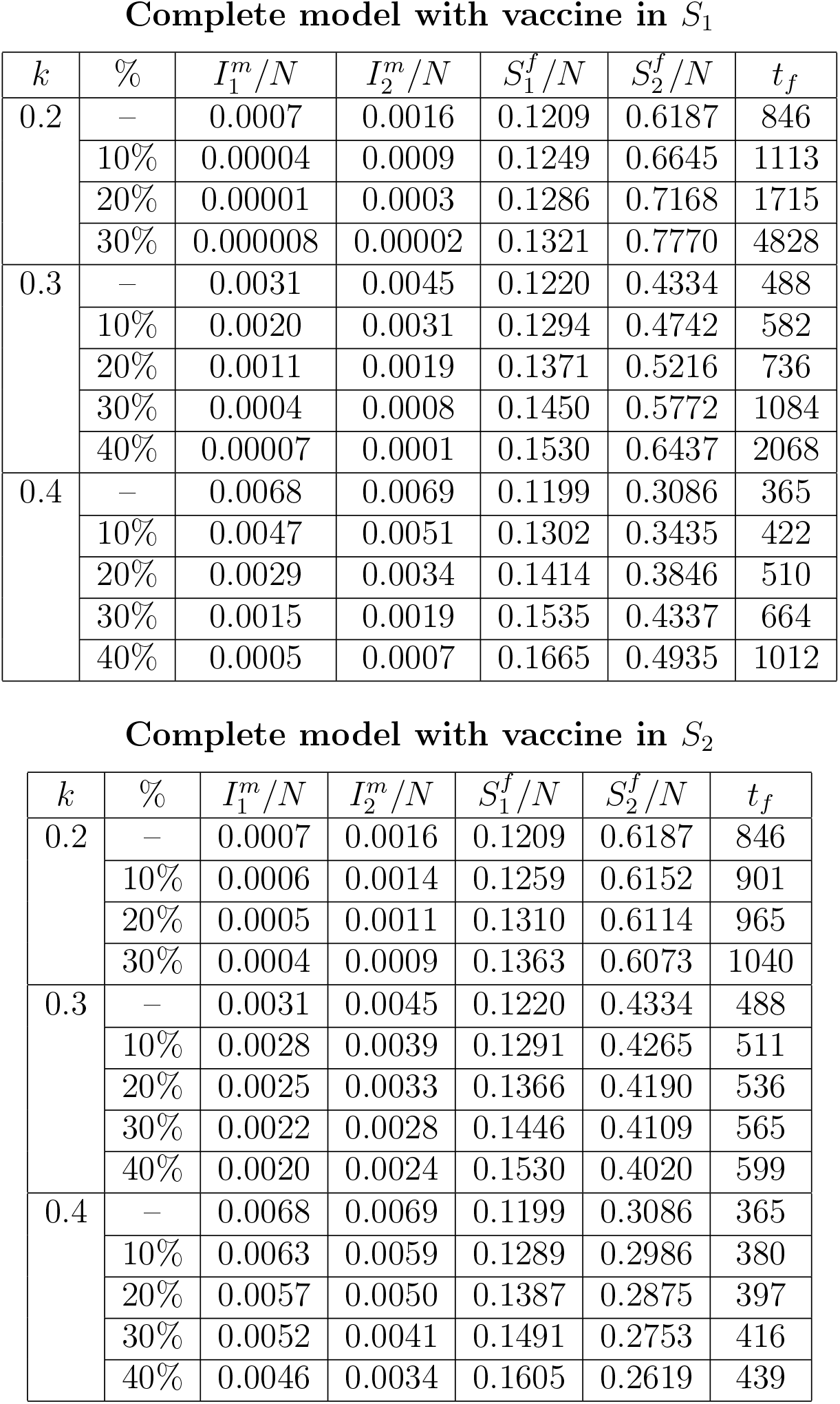
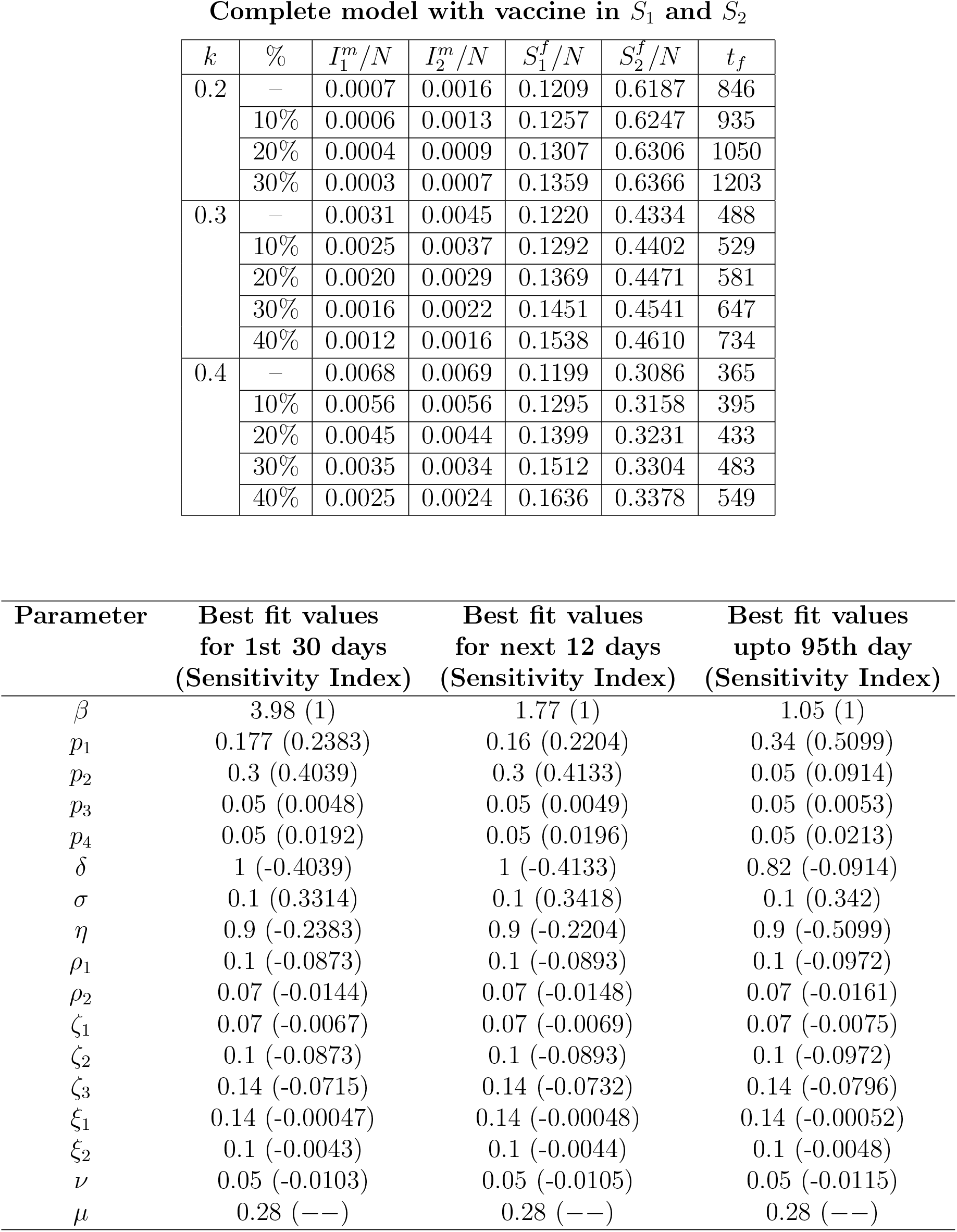

**Complete model with vaccine in $S_1$**
| $k$ | % | $I_1^m/N$ | $I_2^m/N$ | $S_1^f/N$ | $S_2^f/N$ | $t_f$ |
| --- | --- | --- | --- | --- | --- | --- |
| 0.2 | – | 0.0007 | 0.0016 | 0.1209 | 0.6187 | 846 |
|  | 10% | 0.00004 | 0.0009 | 0.1249 | 0.6645 | 1113 |
|  | 20% | 0.00001 | 0.0003 | 0.1286 | 0.7168 | 1715 |
|  | 30% | 0.000008 | 0.00002 | 0.1321 | 0.7770 | 4828 |
| 0.3 | – | 0.0031 | 0.0045 | 0.1220 | 0.4334 | 488 |
|  | 10% | 0.0020 | 0.0031 | 0.1294 | 0.4742 | 582 |
|  | 20% | 0.0011 | 0.0019 | 0.1371 | 0.5216 | 736 |
|  | 30% | 0.0004 | 0.0008 | 0.1450 | 0.5772 | 1084 |
|  | 40% | 0.00007 | 0.0001 | 0.1530 | 0.6437 | 2068 |
| 0.4 | – | 0.0068 | 0.0069 | 0.1199 | 0.3086 | 365 |
|  | 10% | 0.0047 | 0.0051 | 0.1302 | 0.3435 | 422 |
|  | 20% | 0.0029 | 0.0034 | 0.1414 | 0.3846 | 510 |
|  | 30% | 0.0015 | 0.0019 | 0.1535 | 0.4337 | 664 |
|  | 40% | 0.0005 | 0.0007 | 0.1665 | 0.4935 | 1012 |

**Complete model with vaccine in $S_2$**
| $k$ | % | $I_1^m/N$ | $I_2^m/N$ | $S_1^f/N$ | $S_2^f/N$ | $t_f$ |
| --- | --- | --- | --- | --- | --- | --- |
| 0.2 | – | 0.0007 | 0.0016 | 0.1209 | 0.6187 | 846 |
|  | 10% | 0.0006 | 0.0014 | 0.1259 | 0.6152 | 901 |
|  | 20% | 0.0005 | 0.0011 | 0.1310 | 0.6114 | 965 |
|  | 30% | 0.0004 | 0.0009 | 0.1363 | 0.6073 | 1040 |
| 0.3 | – | 0.0031 | 0.0045 | 0.1220 | 0.4334 | 488 |
|  | 10% | 0.0028 | 0.0039 | 0.1291 | 0.4265 | 511 |
|  | 20% | 0.0025 | 0.0033 | 0.1366 | 0.4190 | 536 |
|  | 30% | 0.0022 | 0.0028 | 0.1446 | 0.4109 | 565 |
|  | 40% | 0.0020 | 0.0024 | 0.1530 | 0.4020 | 599 |
| 0.4 | – | 0.0068 | 0.0069 | 0.1199 | 0.3086 | 365 |
|  | 10% | 0.0063 | 0.0059 | 0.1289 | 0.2986 | 380 |
|  | 20% | 0.0057 | 0.0050 | 0.1387 | 0.2875 | 397 |
|  | 30% | 0.0052 | 0.0041 | 0.1491 | 0.2753 | 416 |
|  | 40% | 0.0046 | 0.0034 | 0.1605 | 0.2619 | 439 |

**Complete model with vaccine in $S_1$ and $S_2$**
| $k$ | % | $I_1^m/N$ | $I_2^m/N$ | $S_1^f/N$ | $S_2^f/N$ | $t_f$ |
| --- | --- | --- | --- | --- | --- | --- |
| 0.2 | — | 0.0007 | 0.0016 | 0.1209 | 0.6187 | 846 |
|  | 10% | 0.0006 | 0.0013 | 0.1257 | 0.6247 | 935 |
|  | 20% | 0.0004 | 0.0009 | 0.1307 | 0.6306 | 1050 |
|  | 30% | 0.0003 | 0.0007 | 0.1359 | 0.6366 | 1203 |
| 0.3 | — | 0.0031 | 0.0045 | 0.1220 | 0.4334 | 488 |
|  | 10% | 0.0025 | 0.0037 | 0.1292 | 0.4402 | 529 |
|  | 20% | 0.0020 | 0.0029 | 0.1369 | 0.4471 | 581 |
|  | 30% | 0.0016 | 0.0022 | 0.1451 | 0.4541 | 647 |
|  | 40% | 0.0012 | 0.0016 | 0.1538 | 0.4610 | 734 |
| 0.4 | — | 0.0068 | 0.0069 | 0.1199 | 0.3086 | 365 |
|  | 10% | 0.0056 | 0.0056 | 0.1295 | 0.3158 | 395 |
|  | 20% | 0.0045 | 0.0044 | 0.1399 | 0.3231 | 433 |
|  | 30% | 0.0035 | 0.0034 | 0.1512 | 0.3304 | 483 |
|  | 40% | 0.0025 | 0.0024 | 0.1636 | 0.3378 | 549 |

| Parameter | Best fit values<br>for 1st 30 days<br>(Sensitivity Index) | Best fit values<br>for next 12 days<br>(Sensitivity Index) | Best fit values<br>upto 95th day<br>(Sensitivity Index) |
| --- | --- | --- | --- |
| $\beta$ | 3.98 (1) | 1.77 (1) | 1.05 (1) |
| $p_1$ | 0.177 (0.2383) | 0.16 (0.2204) | 0.34 (0.5099) |
| $p_2$ | 0.3 (0.4039) | 0.3 (0.4133) | 0.05 (0.0914) |
| $p_3$ | 0.05 (0.0048) | 0.05 (0.0049) | 0.05 (0.0053) |
| $p_4$ | 0.05 (0.0192) | 0.05 (0.0196) | 0.05 (0.0213) |
| $\delta$ | 1 (-0.4039) | 1 (-0.4133) | 0.82 (-0.0914) |
| $\sigma$ | 0.1 (0.3314) | 0.1 (0.3418) | 0.1 (0.342) |
| $\eta$ | 0.9 (-0.2383) | 0.9 (-0.2204) | 0.9 (-0.5099) |
| $\rho_1$ | 0.1 (-0.0873) | 0.1 (-0.0893) | 0.1 (-0.0972) |
| $\rho_2$ | 0.07 (-0.0144) | 0.07 (-0.0148) | 0.07 (-0.0161) |
| $\zeta_1$ | 0.07 (-0.0067) | 0.07 (-0.0069) | 0.07 (-0.0075) |
| $\zeta_2$ | 0.1 (-0.0873) | 0.1 (-0.0893) | 0.1 (-0.0972) |
| $\zeta_3$ | 0.14 (-0.0715) | 0.14 (-0.0732) | 0.14 (-0.0796) |
| $\xi_1$ | 0.14 (-0.00047) | 0.14 (-0.00048) | 0.14 (-0.00052) |
| $\xi_2$ | 0.1 (-0.0043) | 0.1 (-0.0044) | 0.1 (-0.0048) |
| $\nu$ | 0.05 (-0.0103) | 0.05 (-0.0105) | 0.05 (-0.0115) |
| $\mu$ | 0.28 (—) | 0.28 (—) | 0.28 (—) |

Best fitted values for the parameters of the full model for Germany [9].

